# Interventions to reduce opioid use for patients with chronic non-cancer pain in primary care settings: a systematic review and meta-analysis

**DOI:** 10.1101/2024.03.13.24304059

**Authors:** Qian Cai, Christos Grigoroglou, Thomas Allen, Teng-Chou Chen, Li-Chia Chen, Evangelos Kontopantelis

## Abstract

**Objective:** This systematic review and meta-analysis aimed to assess interventions to reduce opioid use for patients with chronic non-cancer pain (CNCP) versus usual care or active controls in primary care settings.

**Methods:** In this registered study (PROSPERO: CRD42022338458), we searched MEDLINE, Embase PsycInfo, CINAHL, and Cochrane Library from inception to December 28^th^ 2021, and updated on Dec 14^th^ 2023 for randomized controlled trials (RCTs) and cohort studies with no restrictions. Methodological quality was assessed using the Cochrane Risk of Bias tool for RCTs and Newcastle Ottawa Scale for cohort studies. Primary outcomes included mean reduction in morphine equivalent daily dose (reported as mean differences [MDs] mg/day; 95% confidence intervals [95%CIs]) and/or opioid cessation proportion. Secondary outcomes were mean changes in pain severity (reported as standardized mean difference [SMDs]; 95%CIs) and (serious) adverse events. Meta-analyses were performed using random-effects models.

**Results:** We identified 3,826 records, of which five RCTs (953 participants) and six cohort studies (967 participants) were included. Overall, opioid dosage was significantly reduced in intervention groups compared to controls (MD: -24.88 mg/day, 95%CI: -36.40 to -13.36; I^2^=59.41%; nine studies). Subgroup analyses revealed significant opioid dose reductions with mindfulness (MD: -29.36 mg/day 95%CI: - 40.55 to -18.17; I^2^=0.0%; two trials) and CBT-based multimodalities (MD: -41.68 mg/day; 95%CI: -58.47 to -24.89; I^2^=0.0%; two cohort studies), respectively, compared to usual care. No significant differences were observed in opioid cessation (Odds ratio: 1.55, 95%CI: 0.3 to 2.81, I^2^=50.79%; three studies) or pain severity (SMD: -0.13, 95%CI: -0.37 to 0.11; I^2^=33.51%; three trials). Adverse events were infrequently examined, with withdrawal symptoms commonly reported.

**Conclusions:** The studied interventions were effective in reducing opioid dosage for people with CNCP in primary care. They highlighted the importance of multidisciplinary collaboration. Large-scale RCTs measuring the long-term effects and cost of these interventions are needed before their implementation.

## Introduction

Opioids are primarily recommended by the World Health Organization pain ladder in treating acute pain, cancer pain, and palliative care [1]. During the last two decades, there has been a marked increase in opioid prescriptions issued for chronic non-cancer pain (CNCP), predominantly in the United States (US) [2], Canada [3], and several European countries [4–6] including the United Kingdom (UK) [7]. Despite the widespread utilization of opioids, findings from randomized controlled trials and systematic reviews suggest limited efficacy of these medications, yielding only modest effects in pain relief in the short to medium term (less than 12 weeks) [8]. While the evidence supporting the long-term use of opioids remains sparse, the associated harms of long-term opioid treatment (LTOT), including respiratory depression, bone fractures, and opioid-related mortality, are well documented [9]. Moreover, prolonged opioid use introduces risks of dependence, addiction, and abuse [10].

In response to these concerns, clinical guidelines [11–13] have prompted healthcare providers (HCPs) to reassess their prescribing practices, emphasizing the need to reduce opioid use when potential risks outweigh perceived benefits. However, the endeavor to reduce opioid use encounters significant challenges, including patients’ fear of withdrawal symptoms, inadequate social and healthcare support, and limited availability of non-opioid methods for pain management [14, 15]. Currently, there is a lack of guidance and practical support to implement the reduction of LTOT. Therefore, there is an urgent need for an evidence-based evaluation of interventions to assess their effectiveness in reducing opioid utilization and evaluate their impact on clinical outcomes, particularly in primary care settings, where opioids are mostly prescribed.

Previous systematic reviews [16–19] have provided valuable insights into interventions aimed at reducing opioid use for chronic pain patients. However, their inclusion criteria were broad, limiting their relevance to primary care settings. For example, these reviews analyzed interventions (e.g., spinal cord stimulation) that are not readily accessible in primary care settings, and they included studies that evaluated abrupt or gradual opioid tapering protocols without incorporating additional supportive therapies for patients. The provision of supplementary interventions is crucial, as patients often express reluctance to reduce or cease opioids when alternative treatments are not provided, given these medications constitute their primary method of pain management in real-world practices. Furthermore, the absence of additional interventions impedes the identification of the key components contributing to the dose reduction outcomes. Some prior systematic reviews also included pilot studies lacking clear objectives of opioid reduction. This is particularly common for studies using pharmacological substitution, where the primary aim was to address opioid withdrawal symptoms rather than reducing opioid dosage, despite achieving a decrease in opioid dosage or an improvement in pain severity. In view of these limitations and the emergence of new studies recently, there is a compelling need for an updated evaluation of the current evidence.

The present systematic review and meta-analysis aimed to evaluate the effectiveness of healthcare provider-directed interventions designed to reduce or discontinue opioid use for patients with CNCP, with a particular focus on primary care settings. Specifically, the objectives included comparing changes in morphine equivalent daily dose (MEDD), evaluating the proportion of patients discontinuing opioids, and assessing the change in pain severity and adverse events between interventions and usual care or active controls.

## Methods

This registered systematic review and meta-analysis (PROSPERO: CRD42022338458) was conducted following the Cochrane Handbook [20] and the Preferred Reporting Items for

Systematic Reviews and Meta-Analyses (PRISMA) guidelines (S1 Appendix) [21].

### Eligibility criteria

Eligible studies were full-scale randomized controlled trials (RCTs) and cohort studies that examined primary care provider-directed interventions, designed to reduce or discontinue opioids in adult patients (≥18 years) with CNCP (persists for ≥3 months). Studies that concurrently used opioids with other analgesics (e.g., non-steroidal anti-inflammatory drugs) were excluded, as distinguishing the analgesic effects of opioids from those analgesics were challenging. Interventions implemented in hospital settings or managed by patient themselves were not considered for inclusion. Studies that solely focused on reducing opioids without offering patients alternate treatments or replacements were excluded. Interventions were considered if they explicitly aiming to reduce or cease opioid use, whereas those with a spillover effect on opioid use were excluded. Any comparator was accepted, including usual care or active controls (either pharmacological or non-pharmacological).

The primary outcomes of interest included: 1) opioid dose reduction, measured by the mean changes in morphine milligram equivalent daily dose (MEDD) from pre- to post-treatment. The homogeneity of this outcome measure allowed us to examine our study objectives meta-analytically; 2) The proportion of participants for whom opioid use was either ceased or declined. Secondary outcomes included the mean change in pain severity (measured by pain rating scales on a range of 0-10, where 0 indicating no pain whilst 10 indicating severe pain) and the number of (serious) adverse events related to opioid reduction. Case reports, cross-sectional studies, case-control studies, pilot studies, reviews or meta-analyses were excluded.

### Data sources and search strategies

We performed comprehensive searches in databases including MEDLINE, EMBASE, PsycINFO, CINAHL, and Cochrane Central Register of Controlled Trials. Searches were conducted from the inception of each database until December 28^th^ 2021, and updated on December 14^th^ 2023, using structured search strategies (S2 Appendix) that incorporated text words and medical subject headings related to “chronic pain”, “opioids”, and “reduce/discontinue/cease/deprescribe/”. No language and geographic restrictions were applied. Ongoing trials or unpublished studies were obtained from ClinicalTrials.gov. To ensure literature saturation, we manually retrieved additional studies from the reference lists of included studies and published systematic reviews.

### Study selection

Two review authors (QC and CG) independently screened titles and abstracts of the retrieved citations against pre-determined inclusion/exclusion criteria. When necessary, full text of eligible records were reviewed for further eligibility assessment. Where discrepancies occurred and remained unresolved by discussion, a third party (EK, TA and LCC) was consulted for adjudication. The inter-rater agreement test demonstrated a high level of consistency (99.64%) between QC and CG with a Kappa coefficient equals to 0.7982 (p<0.001).

### Data extraction and quality assessment

Data extraction was undertaken independently by one review author (QC) and verified by another reviewer (TCC) for accuracy. The study authors were also contacted by email to request any necessary missing information. The following key information was extracted:

- Study: first author, publication year, country, study design, settings.
- Patient characteristics: age, gender, sample size, pain duration and severity, opioid use status, comorbidities.
- Intervention and comparator: key components, mode of administration, frequency, treatment duration.
- Outcome: opioid dose, pain severity, adverse events.

### Risk of bias assessment

The methodological quality of all included studies was independently appraised by QC. For RCTs, the Cohrane’s Risk of Bias Tool 2.0 (RoB 2) [22] was employed to assess the risk of bias, including 1. sequence generation; 2. allocation concealment; 3. masking of participants, staff and outcome assessors; 4. incomplete outcome data; and 5. selective outcome reporting. For cohort studies, a modified Newcastle-Ottawa Scale (NOS) [23] was used to evaluate bias risk, focusing on 1. the representativeness of the exposed cohort; 2. selection of the non-exposed cohort; 3. ascertainment of exposure; 4. absence of the outcome of interest at the start of the study; 5. comparable controls; 6. assessment of outcome; 7. sufficient follow-up length for outcomes to occur; and 8. adequacy of follow-up cohorts. For RCTs, high quality was defined as minimum 4 domains of low risk. As for cohort studies, the modified NOS adopted a star rating system. A study was deemed to be of good quality if it obtained ≥3 stars in selection domain (1-4), ≥1 star in comparability domain (5), and ≥2 stars in outcome/exposure domain (6-8).

## Data synthesis and analysis

The study characteristics and details of opioid reduction interventions were presented descriptively. Effect sizes were reported using odds ratios (ORs) with their 95% confidence intervals (95%CIs) for opioid cessation, mean differences (MDs) with 95%CIs for opioid dose reduction, and standardized mean differences (SMDs) with 95%CIs for pain severity, given the utilization of different scales for this outcome measure. Magnitude of effect was defined as large (SMD > 0.8), medium (SMD 0.5-0.8), small (SMD 0.2-0.5) or trivial (SMD < 0.2) [24].

Where between- or within-group SDs were not reported, relevant data such as sample size, p values, t statistics, standard errors (SEs), or 95% CIs were used to derive SDs using the formula recommended by the Cochrane Handbook (6.5.2.3) [20] or the calculator embedded in Review Manager 5.4. Additionally, when outcomes were assessed at multiple time points during long-term follow-up, data from the last available time point were employed.

Heterogeneity was assessed using the I^2^ statistics, with values below 50% suggesting low heterogeneity, 50-75% moderate, above 75% substantial heterogeneity [25]. Random effects meta-analyses with a non-parametric bootstrap of DerSimonian Laird (DL) method were conducted for pooling the outcomes of interest [26, 27] even if I^2^ was low. Publication bias regarding the primary outcomes was not assessed due to the small number of included studies.

Sensitivity analyses were conducted by sequentially removing one study at a time and repeating the meta-analysis based on the remaining data to assess whether pooled estimates were unduly influenced by specific studies. Furthermore, multivariable random effects meta-regressions were conducted to explore potential sources of heterogeneity, including patient mean age; gender; study period; sample size; follow-up time point; intervention type and category. Subgroup analyses were undertaken based on intervention types, which included mindfulness techniques, and CBT-based multi-component strategies. All data analyses were conducted with Stata/MP 17.0.

## Results

### Study selection and patient characteristics

The initial search retrieved 3,826 potentially relevant records, of which 1,126 duplicates were removed, yielding 2,770 unique records for eligibility assessment. By screening titles and abstracts against our predefined inclusion/exclusion criteria, 2739 studies were excluded. Of the remaining 31 records, eleven full-text articles including five RCTs (953 participants, female 557 [58.4%], mean age: 60.1±13.42) [28–32] and six retrospective cohort studies (patients 967, female 214 [22.1%], mean age: 54.2±13.52) [33–38] published between 2016 and 2023 were included in this systematic review and meta-analysis (Fig 1. Selection of included studies). All included studies reported the baseline opioid dosage (mean 87.4±131.8 mg/day), with three [31, 33, 37] indicating that participants consumed a mean MEDD of >120 mg/day. Most studies did not specify the exact CNCP conditions, except for one study that explicitly mentioned chronic musculoskeletal pain [38]. Studies were mainly conducted in the US (n=10), with one in the UK [30]. Both RCTs and retrospective cohort studies had a small sample size ranging between 35 and 608 (Table 1).

**Table 1.**
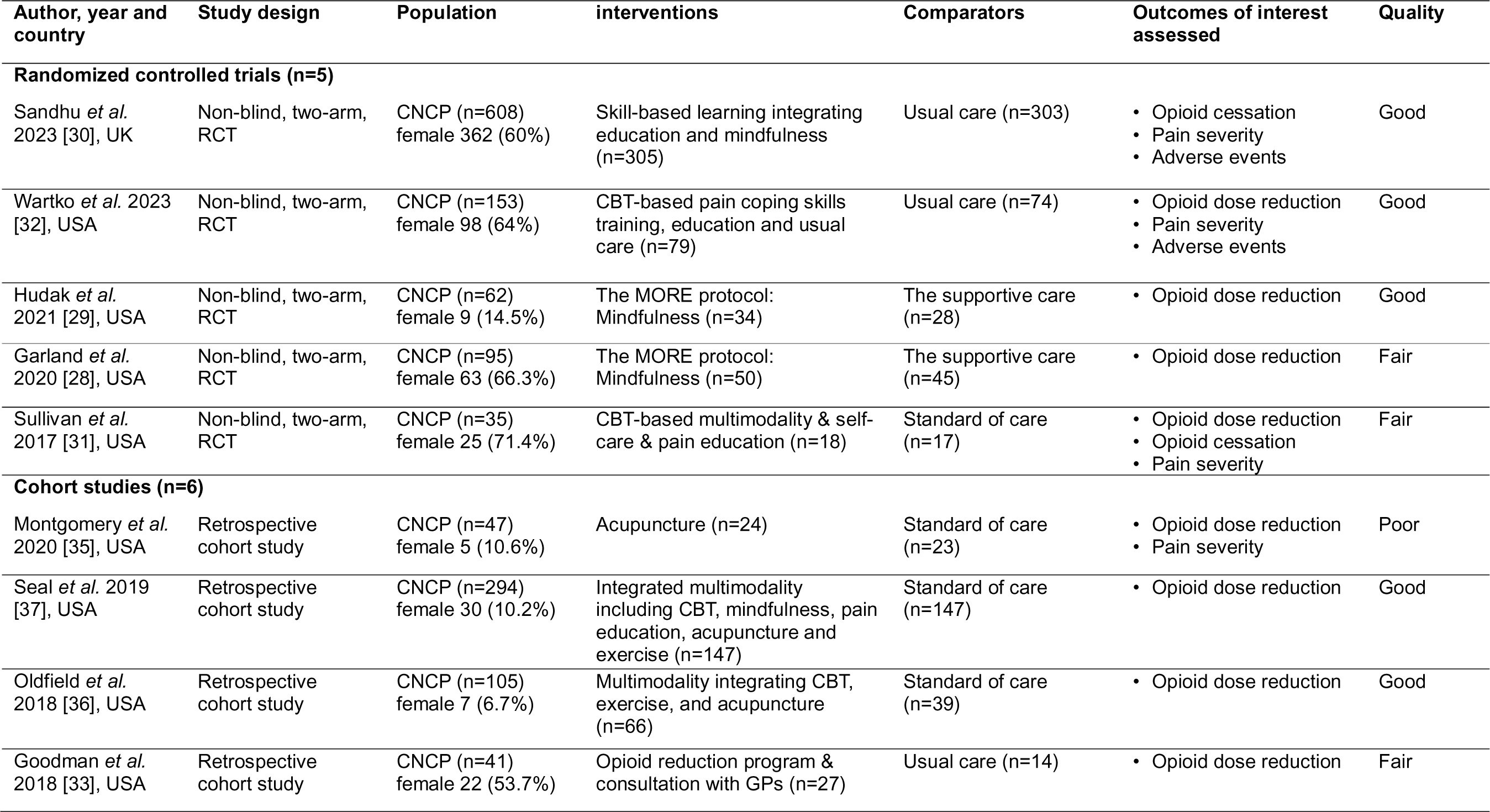
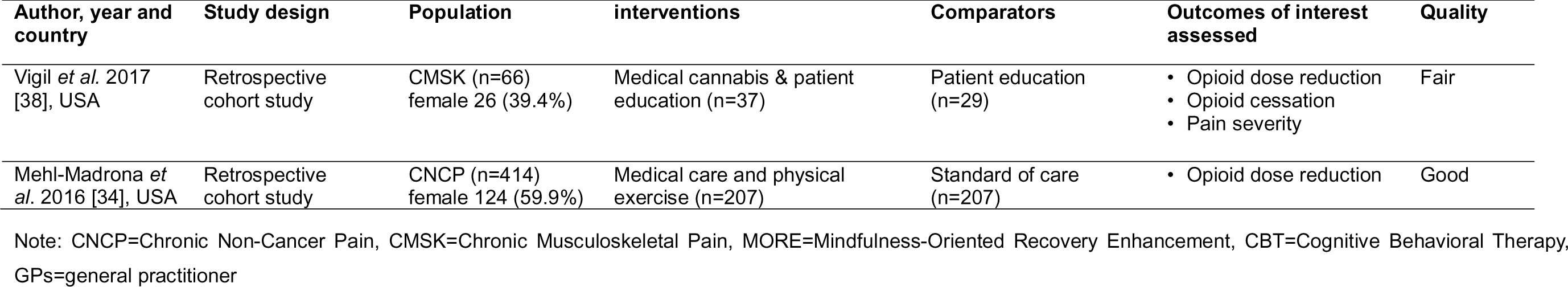
Characteristics of included studies.

| Author, year and country | Study design | Population | interventions | Comparators | Outcomes of interest assessed | Quality |
| --- | --- | --- | --- | --- | --- | --- |
| **Randomized controlled trials (n=5)** |  |  |  |  |  |  |
| Sandhu *et al.* 2023 [30], UK | Non-blind, two-arm, RCT | CNCP (n=608)<br>female 362 (60%) | Skill-based learning integrating education and mindfulness (n=305) | Usual care (n=303) | • Opioid cessation<br>• Pain severity<br>• Adverse events | Good |
| Wartko *et al.* 2023 [32], USA | Non-blind, two-arm, RCT | CNCP (n=153)<br>female 98 (64%) | CBT-based pain coping skills training, education and usual care (n=79) | Usual care (n=74) | • Opioid dose reduction<br>• Pain severity<br>• Adverse events | Good |
| Hudak *et al.* 2021 [29], USA | Non-blind, two-arm, RCT | CNCP (n=62)<br>female 9 (14.5%) | The MORE protocol: Mindfulness (n=34) | The supportive care (n=28) | • Opioid dose reduction | Good |
| Garland *et al.* 2020 [28], USA | Non-blind, two-arm, RCT | CNCP (n=95)<br>female 63 (66.3%) | The MORE protocol: Mindfulness (n=50) | The supportive care (n=45) | • Opioid dose reduction | Fair |
| Sullivan *et al.* 2017 [31], USA | Non-blind, two-arm, RCT | CNCP (n=35)<br>female 25 (71.4%) | CBT-based multimodality & self-care & pain education (n=18) | Standard of care (n=17) | • Opioid dose reduction<br>• Opioid cessation<br>• Pain severity | Fair |
| **Cohort studies (n=6)** |  |  |  |  |  |  |
| Montgomery *et al.* 2020 [35], USA | Retrospective cohort study | CNCP (n=47)<br>female 5 (10.6%) | Acupuncture (n=24) | Standard of care (n=23) | • Opioid dose reduction<br>• Pain severity | Poor |
| Seal *et al.* 2019 [37], USA | Retrospective cohort study | CNCP (n=294)<br>female 30 (10.2%) | Integrated multimodality including CBT, mindfulness, pain education, acupuncture and exercise (n=147) | Standard of care (n=147) | • Opioid dose reduction | Good |
| Oldfield *et al.* 2018 [36], USA | Retrospective cohort study | CNCP (n=105)<br>female 7 (6.7%) | Multimodality integrating CBT, exercise, and acupuncture (n=66) | Standard of care (n=39) | • Opioid dose reduction | Good |
| Goodman *et al.* 2018 [33], USA | Retrospective cohort study | CNCP (n=41)<br>female 22 (53.7%) | Opioid reduction program & consultation with GPs (n=27) | Usual care (n=14) | • Opioid dose reduction | Fair |
| Vigil *et al.* 2017 [38], USA | Retrospective cohort study | CMSK (n=66)<br>female 26 (39.4%) | Medical cannabis & patient education (n=37) | Patient education (n=29) | • Opioid dose reduction<br>• Opioid cessation<br>• Pain severity | Fair |
| Mehl-Madrona *et al.* 2016 [34], USA | Retrospective cohort study | CNCP (n=414)<br>female 124 (59.9%) | Medical care and physical exercise (n=207) | Standard of care (n=207) | • Opioid dose reduction | Good |

Note: CNCP=Chronic Non-Cancer Pain, CMSK=Chronic Musculoskeletal Pain, MORE=Mindfulness-Oriented Recovery Enhancement, CBT=Cognitive Behavioral Therapy, GPs=general practitioner

### Characteristics of interventions

The components, duration, frequency, and delivery mode of the interventions varied significantly across studies. One study used medicine substitution approach, transitioning opioids to cannabis [38]; Two studies focused on physical interventions, incorporating components such as yoga, Taichi, chiropractic therapy [34] and acupuncture [35]; Five studies involved psychological or behavioral changes, integreting key components such as cognitive behavior therapy (CBT), mindfulness techniques, patient education and pain coping skills training [28–30, 32, 33]; Three studies [31, 36, 37] employed mixed multimodal approaches delivered by a multidisciplinary team comprising physicians, nurses, psychologists, pharmacists, and social workers. These multimodal care programs typically combined CBT, mindfulness, acupuncture, chiropractic care, exercise, pharmacotherapies are the core components.

The most common comparator was usual care, with the exception of one study that used patient education as the comparison group [38]. Treatment durations ranged from eight weeks to 12 months. Follow-up periods (median 6 months) varied from short (≤3 months) [28, 29] to intermediate (6-9 months) [31, 33–37], with three studies reporting long-term outcomes of opioid reduction (≥12 months) . (See Table 2 for characteristics of interventions).

**Table 2.**
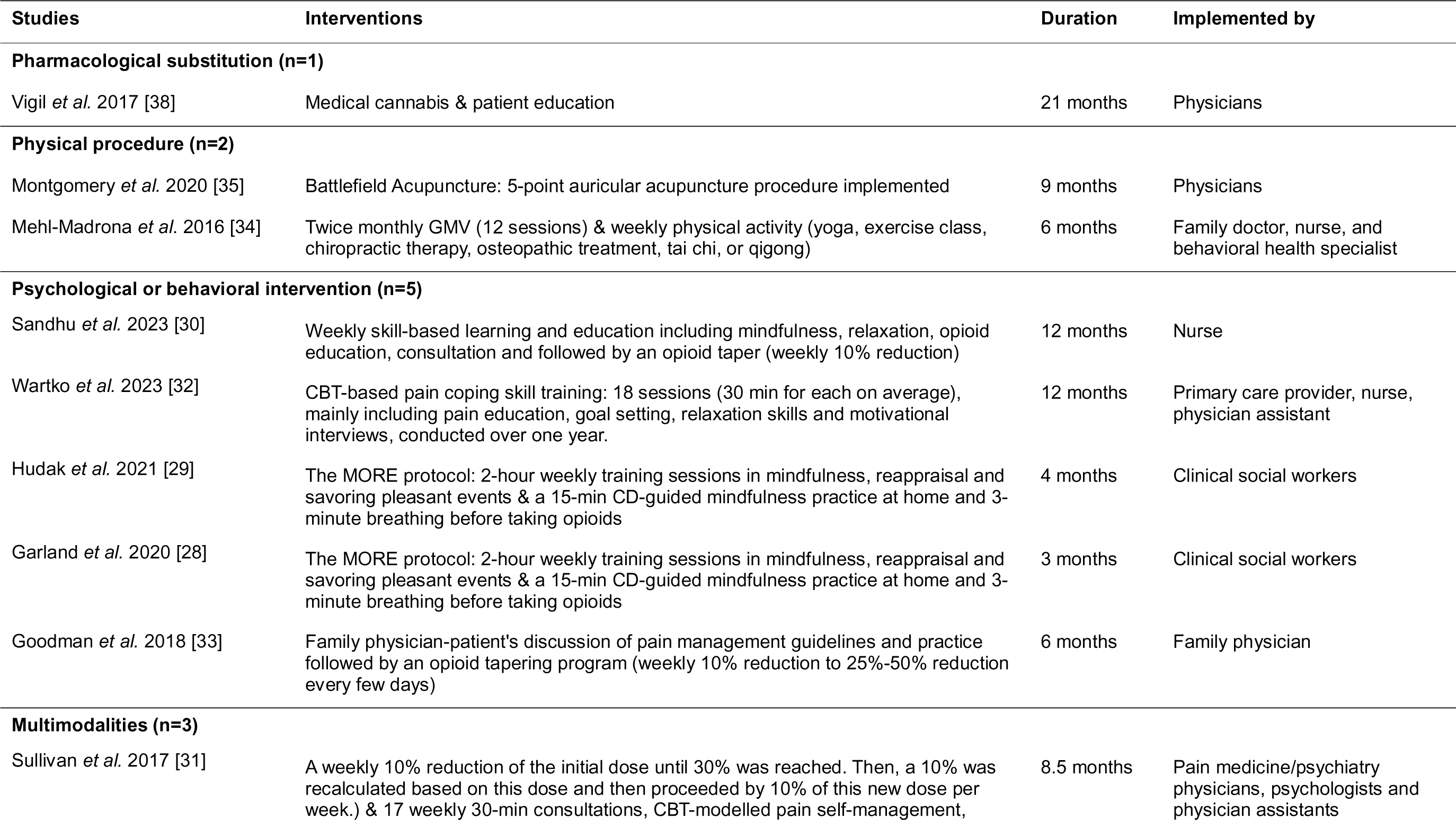
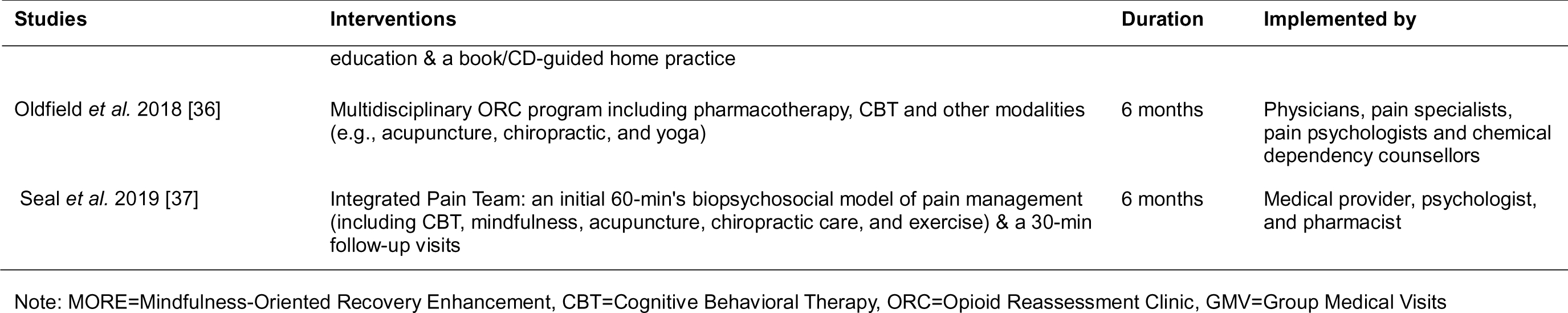
The components and implementation procedures of each intervention.

### Risk of bias assessment

The Cochrane RoB-2 assessment indicated an overall risk of bias being low (n=3) to moderate (n=2) (Fig 2. Risk of Bias of included RCTs). Due to the impossibility of masking the interventions, participants and researchers were unblinded, which increased risk. The high dropout rate during follow-ups was another main reason contributing to the increased risk of bias. The NOS evaluation tool identified three cohort studies [34, 36, 37] with good quality, two [33, 38] with fair and one [35] with poor quality (Table 3). Uncontrolled confounders and limited representativeness of the study population were the main reasons that reduced the quality of these studies.

**Table 3.** Quality assessment outcomes of cohort studies.

| Author and year | Three domains of the Newcastle-Ottawa Scale |  |  | Quality |
| --- | --- | --- | --- | --- |
|  | Selection of participants | Comparability of study groups | Outcomes |  |
| Montgomery *et al.* 2020 [35] | ★ ★ ★ | ★ | ★ | Poor |
| Seal *et al.* 2019 [37] | ★ ★ ★ ★ | ★ ★ | ★ ★ ★ | Good |
| Oldfield *et al.* 2018 [36] | ★ ★ ★ | ★ ★ | ★ ★ ★ | Good |
| Goodman *et al.* 2018 [33] | ★ ★ ★ | ★ | ★ ★ | Fair |
| Vigil *et al.* 2017 [38] | ★ ★ ★ | ★ | ★ ★ | Fair |
| Mehl-Madrona *et al.* 2016 [34] | ★ ★ ★ | ★ ★ | ★ ★ ★ | Good |

### Opioid dose reduction

Ten studies including four RCTs [28, 29, 31, 32] and six cohort studies [33–38] reported the outcomes of opioid dose reduction. Opioid dosage was significantly decreased in the intervention groups compared to controls (MD: -31.80 mg/day, 95%CI: -50.30 to - 13.31; ten studies; Fig 3. Forest plot of interventions vs controls in opioid dose reduction in ten studies). However, considerable heterogeneity was noted (I^2^ = 91.14%, 95%CI: 80.4% to 96.0%, p=0.00; Fig 3). A sensitivity analysis was conducted to investigate the source of heterogeneity and the Mehl-Madrone et al. (2016)’s study [34] was identified as an outlier (Fig 4. Sensitivity analysis of included studies). Upon its removal, the heterogeneity decreased from high (I^2^ = 91.14%) to medium (I^2^ = 59.41%), but the significance or direction of the pooled effects in our meta-analyses remained unchanged (MD: -24.88 mg; 95%CI: -36.40 to -13.36; nine studies; Fig 5. Forest plot of interventions vs controls in opioid dose reduction in nine studies). Furthermore, a multivariable meta-regression analysis was conducted, revealing that heterogeneity could be partially explained by differences in the longest follow-up time across studies (p=0.014).

Within the four RCTs [28, 29, 31, 32] provided data that enabled the pooled calculation of change for opioid reduction among 181 participants receiving different interventions compared to 162 receiving usual care (MD: -24.40 mg; 95%CI: -36.32 to -12.47; I^2^ = 9.21%, p=0.35; Fig 6. Forest plot of interventions vs controls in opioid dose reduction in four RCTs), a subgroup meta-analysis of two trials [28, 29] using the MORE protocol (mindfulness) found a significant reduction in opioid dosage in 84 patients receiving this intervention, compared to 77 having supportive care (MD: -29.36 mg; 95%CI: -40.55 to -18.17; I^2^ = 0.00%, p=0.37; Fig 6). Similar decreases in opioid dose were reported by Wartko et al. (2023) [32] and Sullivan et al. (2017) [31] using CBT-based multi- component interventions. However, no between-group statistical significance was attained (MD: -10.10 mg; 95%CI: -33.61 to 13.40; I^2^ = 0.00%, p=0.52; Fig 6).

Within the six cohort studies [33–38], where various interventions were implemented, a significant decrease in the use of opioid medications was observed in the intervention groups (MD: -29.89 mg; 95%CI: -49.00 to -10.78; I^2^ = 72.72%, p=0.01; Fig 7. Forest plot of interventions vs controls in opioid dose reduction in five cohort studies). Subgroup analysis of Oldfield et al. (2018) and Seal et al. (2021) using CBT-based interventions showed a significant reduction in opioid dose, compared to those in the control group. (MD: -41.68 mg; 95%CI: -58.47 to -24.89; I^2^ = 0.00%, p=0.76; Fig 7).

### Opioid cessation

No significant differences were observed in opioid cessation (Odds ratio: 1.55, 95%CI: 0.3 to 2.81; I2=50.79%, p=0.13; three studies; Fig 8. Forest plot of interventions vs controls in opioid cessation) between intervention groups and controls. In the two RCTs [30, 31], although the interventions had slight variations, both studies implemented a weekly 10% dose reduction protocol combined with education and consultation as core components of the interventions. In the Sandhu et al. (2023) study [30], patients undergoing skill-based learning and education had a significantly higher cessation rate than those receiving usual care (Odds radio [OR]: 5.23; 95% CI 2.87 to 9.52; Fig 8.). However, pooled meta-analysis showed no statistically significant change in opioid discontinuation (OR: 1.10; 95%CI -0.48 to 2.67; Fig 8.). Moderate heterogeneity (I^2^=58.59%, 95% CI 0% to 86.2%) was observed, primarily attributed to the small sample size of the Sullivan et al. (2017) study [31].

In the cohort study [38] reporting opioid cessation, where 37 patients received cannabis-assisted opioid replacement along with patient education. Among them, 15 patients completely discontinued the opioid use over a period of 21 months. When compared to a control group receiving education alone, the cessation rate of opioid was lower in the control group, with only 1 out of 29 patients achieving opioid cessation; however, this difference was not statistically significant, primarily because of the very small sample size (OR: 2.95, 95% CI 0.85 to 5.05; Fig 8).

### Pain severity

Pain severity changes at different observation time points were reported in three trials [30–32], all utilizing interventions grounded in behavior change and patient education. Due to the variations in pain severity measurements across studies, standard mean differences were used for pooling the estimates. While a decrease in pain severity was observed in all three trials, the between-group difference was small and non-significant (SMD -0.13; 95% CI -0.37 to 0.11, I^2^ =33.51%, p=0.22; Fig 9. Forest plot of interventions vs controls in pain severity improvement). It is worth noting that this 0.13- point reduction in pain score might not have meaningful clinical implications.

Due to the poor study quality and high heterogeneity, meta-analysis was not feasible for two cohort studies [35, 38] reporting the outcome of pain severity. Therefore, the results were narratively reviewed. Compared with these behavioral, psychological and physical interventions, the pharmacological intervention (cannabis) combined with patient education, as used in the Vigil et al. (2017) study [38], showed a significant decrease in pain intensity score (MD: -3.4 on a scale of 0-10, p<0.001), which suggested a meaningful clinical implication. Acupuncture, as employed in the Montgomery et al. (2020) study [35], exhibited an immediate effect in alleviating pain (a reduction in pain intensity score of MD 1.3 on a scale of 0-10, p<0.01). However, there were no significant differences after 9 months (p = 0.15), indicating acupuncture’s short-term effects.

### Adverse events

Adverse events (AEs) were infrequently examined in included studies, with the majority of reported AEs being associated with psychological and nervous system effects. In Wartko et al. (2023)’s study [32], which involved 79 participants receiving CBT-based pain coping skills training plus usual care, six cases of increased pain, one case of withdrawal symptoms, and one case of anxiety were noted. No serious adverse events (SAEs) were documented in this study. In Sandhu et al. (2023) [30], adverse events such as sleep disturbance, suicidal ideation, headache, withdrawal symptoms were reported by 22 of 305 participants (7%) and 8 of 303 participants (3%), in the intervention and usual care groups, respectively. SAEs occurred in 8% (25/305) of the participants in the intervention group and 5% (16/303) in the usual care group, with the most common SAEs being gastrointestinal disorder, metastatic cancer, hospitalization due to increased pain, and unknown cause of death.

## Discussion

### Summary of main findings

In this systematic review and meta-analysis, we examined 11 studies (five RCTs and six cohort studies) assessing the effectiveness of opioid reduction interventions for patients with CNCP in primary healthcare settings. Despite methodological differences, both RCTs and observational studies yielded statistically similar results, allowing for the pooling of results across the full range of eligible studies. Overall, intervention groups exhibited significant reductions in opioid dosage compared to usual care or active control. Subgroup analyses revealed significant opioid dose reductions with mindfulness and CBT-based multimodalities, respectively, compared to usual care. No significant differences were observed in opioid cessation or pain severity. Adverse events were infrequently examined, with withdrawal symptoms commonly reported. Medium heterogeneity was observed across all studies for each outcome, likely stemming from variability in interventions, follow-up durations, and healthcare providers involved. Nevertheless, the I^2^ values for subgroup analyses were low.

### Compare with existing literature

Our study conclusion generally aligns with previous systematic reviews [16–19] that the strength of evidence was insufficient to draw conclusions. However, our study extends these reviews by providing an updated and comprehensive assessment of the literature, adding one recent good-quality RCT. Supported by subgroup meta-analyses, we elucidated the effectiveness of mindfulness in reducing LTOT. Meanwhile, we highlighted key components such as CBT, patient education, mindfulness, exercise, which may contribute to the successful opioid reduction and thus merit consideration for future intervention development. Importantly, our study exclusively targeted primary care settings, distinguishing it from others that included secondary and tertiary care settings. Although de Kleijn et al. (2021) [17] solely focused on primary care settings, their results were narratively synthesized, and they included studies using simple opioid tapering protocols or interventions targeted at physicians.

Clinical guidelines [11, 12] recommend an initial reduction of opioid dosage by 10% per week, with adjustments for high-dose users (> MEDD 120 mg/day) or those on LTOT exceeding one year [39]. In our study, we observed a pooled mean opioid reduction of 24.88 mg/day from a baseline of 87.4 mg/day. Although this reduction was aggressive and exceeded the guideline-recommended reduction rate, it did yield a clinically meaningful decrease in opioid dosage without worsening pain severity. It is important to acknowledge that adherence to guideline recommendations does not necessarily guarantee optimal patient outcomes, as individual responses to opioid reduction strategies may vary. In fact, the primary goal of CNCP management is to maintain body function rather than achieve complete pain eradiation. Therefore, efforts should prioritize reducing opioids to minimize potential harms associated with LTOT. Meanwhile, despite limited examination of adverse events in this study, our findings align with existing evidence [40, 41], suggesting that opioid reduction may lead to withdrawal symptoms, increased pain severity, suicides or all-cause mortality. Therefore, close monitoring of patient responses to opioid reduction strategies is essential for optimizing treatment outcomes while mitigating adverse effects.

Our findings highlighted the effectiveness of the MORE protocol, with mindfulness as its core component, in reducing opioid dose for CNCP patients in the short term (3 months). Additionally, meta-analytic results indicated a significant and strong association between mindfulness and pain reduction [42]. In another study [43] conducted by the same author using the MORE protocol, participants reported significantly greater improvements in psychological and physical function outcomes (e.g., general activity, mood, walking ability, normal work, relationships, sleep, and enjoyment of life.) compared to the support group during the three-month follow-up period. In view of this evidence, mindfulness could be considered a potential component in the development of opioid reduction strategies. However, before recommending widespread implementation of this intervention, further validation through replication in a second trial with a longer observation period (≥12 months) within primary care settings is necessary.

Contrary to expectations, CBT-based interventions did not show a significant reduction in opioid dosage in RCTs [31, 32]. This finding aligns with recent evidence from a recent cluster RCT, indicating that although CBT significantly reduced pain severity, it did not result in significant differences in opioid use between groups [43]. The reason for this non-significant result may be twofold. First, the small sample size and variations in interventions across studies may have contributed to the lack of statistical significance. Second, the study conducted by Sullivan et al. (2017) [31] involved high- dose users (207±245 mg/day), suggesting that behavior changes might not be superior to usual care in reducing opioids dosage for this population. However, the pain relief achieved through CBT may contribute to a subsequent decrease in opioid utilization, although this effect may take time to manifest. Two observational studies [36, 37] employing similar CBT-based interventions reported significant reductions in opioid dosage compared to usual care. This inconsistency with findings from RCTs may be attributed to the variations in interventions. Additionally, observational studies may yield more promising results due to unadjusted confounders. Further investigation with long- term follow-up and larger sample sizes is warranted to elucidate the effects of CBT on opioid reduction.

Pharmacological interventions (cannabis-assisted opioid substitution) showed promising result, with a higher rate of opioid cessation and improved pain severity observed among the study population. This could be explained by the fact that patients often perceive pharmacotherapies as providing immediate pain relief, contrasting with non-pharmacological approaches which may require more time to yield noticeable effects. Consequently, patients may be more inclined towards pharmacological treatments, especially during episodes of exacerbated pain. This preference may be influenced by patients’ desire to promptly alleviate pain [44], and the challenges associated with accessing non-pharmacological therapies [45], which further perpetuates the reliance on pharmacotherapy in managing CNCP symptoms. While cannabis demonstrates efficacy in replacing opioids for CNCP management, its long- term use may pose potential risks, including lower educational achievement, psychiatric illness, and residual cognitive impairment [46]. Therefore, careful consideration and monitoring are necessary when utilizing cannabis as a substitute for opioids in CNCP management.

Current clinical guidelines from the UK NICE (National Institute for Health and Care Excellence) [13, 47] and NHS Oxford Hospital [12] advocate for a holistic approach beyond pharmacotherapy alone. Our research findings align with these recommendations, revealing that a combination of cognitive change strategies, acupuncture, exercise, patient education and pharmacotherapies, delivered by a multidisciplinary team were more effective in opioid reduction. Central to this success is the practice of shared decision making between patient and healthcare professionals. The collaboration among diverse healthcare professionals (GPs, nurses, therapists, pharmacists, etc) is crucial, as each contributes unique expertise and insights to the table. Nevertheless, challenges remain in accessing to multimodal cares, particularly for individuals unable to take time off work, those residing in regions with limited services, and individuals from culturally and linguistically diverse communities. Addressing these access barriers is essential to ensuring equitable healthcare delivery.

### Strengths and limitations

To our knowledge, this is the first and most up-to-date systematic review and meta- analysis, to evaluate the effectiveness of interventions on opioid dose reduction and pain outcomes for patients with CNCP, with a particular emphasis on primary healthcare settings. By conducting a comprehensive literature search and including recently conducted studies in the UK, this research holds informative value for the potential implementation of interventions outside the US primary care settings. Moreover, several key subgroup analyses were conducted to identify which intervention (components) are effective in achieving opioid reduction purpose.

Our study also had several limitations. First, the relatively small number of included participants, short follow-up periods, and lack of blinding to interventions may have introduced biases and impacted the overall quality of the included studies. Second, data in observational studies were sourced from secondary databases or electronic medical records, which could cause information bias. Third, funnel plots and the Egger’s test were not performed due to the small number of studies included in the meta-analyses Fourth, the majority of the studies were conducted on white populations in the US, likely influenced by the opioid crisis in that country, thus limited the generalizability of our research findings to other ethnicities and regions. Lastly, data was insufficient to support meta-analyses of quality of life or function.

## Conclusion

Although according to prior studies, no specific interventions can be recommended over one another, multidisciplinary opioid reduction strategies, incorporating components such as CBT, pain education, mindfulness, exercise and acupuncture, have demonstrated effectiveness and tolerability in reducing opioid doses and improving pain severity among patients with CNCP in primary care settings. However, robust evidence remains limited due to several factors, including a shortage of well- designed research studies, inadequate long-term follow-up periods and absence of QoL-related outcomes. Future studies should prioritize investigating the active components within these interventions to gain a deeper understanding of the specific strategies contributing to positive outcomes. Additionally, it is essential to assess the cost-effectiveness and patient acceptability of these interventions before their implementation in specific primary care contexts.

## Data Availability

All relevant data are within the manuscript and its Supporting Information files.

## Acknowledgments

None.

## Contributors

Conceptualization: LCC, EK, QC, TCC.

Data curation: QC, CG, TCC, EK.

Formal analysis: QC, EK.

Investigation: QC, CG,TCC.

Methodology: QC, EK.

Project administration: QC.

Supervision: EK TA CG LCC.

Validation: CG TCC.

Visualization: QC.

Writing – original draft: QC.

Writing – review & editing: QC CG TA TCC LCC EK.

