## Supplementary material for "Interventions to reduce opioid use for patients with chronic non-cancer pain in primary care settings: a systematic review and meta-analysis": S1 Appendix

Appendix 1.	Search strategies on the five medical databases

**Ovid MEDLINE**

| 1 | Chronic Pain/ |
| --- | --- |
| 2 | (chronic adj4 pain*).mp. [mp=title, abstract, heading word, drug trade name, original title, device manufacturer, drug manufacturer, device trade name, keyword heading word, floating subheading word, candidate term word] |
| 3 | exp Osteoarthritis/ |
| 4 | osteo-arthritis.mp. |
| 5 | osteoarthr*.mp. [mp=title, abstract, heading word, drug trade name, original title, device manufacturer, drug manufacturer, device trade name, keyword heading word, floating subheading word, candidate term word] |
| 6 | degenerative arthrit*.mp. |
| 7 | exp rheumatoid arthritis/ |
| 8 | osteo?arthrit*.mp. |
| 9 | exp Neuralgia/ |
| 10 | Diabetic Neuropathies.mp. or Diabetic Neuropathies/ |
| 11 | (neuropath* adj5 (pain* or diabet*)).mp. |
| 12 | Neuralgia/ or neuralg*.mp. |
| 13 | fibromyalg*.mp. |
| 14 | ((noncancer* or non-cancer*or chronic* or recurrent or persist* or non-malign* or long term or longterm or wide spread or wide spread chronic) adj3 pain).mp. |
| 15 | (complex regional pain syndromes or causalgia).mp. |
| 16 | Hyperalgesia/ |
| 17 | Trigeminal Neuralgia/ or nerve neuralgia.mp. |
| 18 | (acute or emergency or cancer pain or pr preoperative or postoperative or palliative care or surgical).ti,ab. |
| 19 | deprescrib*.mp. |
| 20 | de-prescrib*.mp. |
| 21 | deprescription*.mp. |
| 22 | de-prescription*.mp. |
| 23 | discontin*.mp. |
| 24 | taper*.mp. |
| 25 | wean*.mp. |
| 26 | switch*.mp. |
| 27 | cess*.mp. |
| 28 | terminat*.mp. |
| 29 | substitut*.mp. |
| 30 | reflex sympathetic dystrophy.mp. |
| 31 | phantom limb.mp. or agnosia/ or phantom pain/ or amputation stump/ |
| 32 | exp backache/ |
| 33 | radiculopathy.mp. or exp radiculopathy/ |
| 34 | musculoskeletal pain/ |
| 35 | exp arthralgia/ |
| 36 | headache/ |
| 37 | headache*.mp. |
| 38 | exp migraine/ |
| 39 | migraine.mp. |
| 40 | fibromyalgia/ |
| 42 | exp narcotic analgesic agent/ |
| 43 | (opioid* or opiate*).mp. |
| 44 | (alfentanil or alphaprodine or beta-casomorphin$ or buprenorphine or carfentanil or codeine or deltorphin or dextromethorphan or dezocine or dihydrocodeine or dihydromorphine or enkephalin$ or ethylketocyclazocine or ethylmorphine or etorphine or fentanyl or heroin or hydrocodone or hydromorphone or ketobemidone or levorphanol or lofentanil or meperidine or meptazinol or methadone or methadyl acetate or morphine or nalbuphine or opium or oxycodone or oxymorphone or pentazocine or phenazocine or phenoperidine or pirinitramide or promedol or propoxyphene or remifentanil or sufentanil or tilidine or tapentadol).mp. |
| 45 | (adolonta or Anpec or Ardinex or Asimadoline or Alvimopam or amadol or biodalgic or biokanol or Codinovo or contramal or Demerol or Dicodid or Dihydrocodeinone or dihydromorphinone or dihydrohydroxycodeinone or dihydrone or dilaudid or dinarkon or dolsin or dolosal or dolin or dolantin or dolargan or dolcontral or duramorph or duromorph or duragesic or durogesic or eucodal or Fedotzine or Fentanest or Fentora or Fortral or Hycodan or Hycon or Hydrocodone or Hydrocodeinonebitartrate or hydromorphon or hydroxycodeinon or isocodeine or isonipecain or jutadol or laudacon or l dromoran or levodroman or levorphan or levo-dromoran or levodromoran or lexir or lidol or lydol or morfin or morfine or morphia or morphin or morphinium or morphinene or morphium or ms contin or n-methylmorphine or n methylmorphine or nobligan or numorphan or oramorph or oxycodeinon or oxiconum or oxycone or oxycontin or palladone or pancodine or pethidine or phentanyl or prontofort or robidone or skenan or sublimaze or sulfentanyl or sulfentanil or sufenta or takadol or talwin or theocodin or tramadol or tramadolhameln or tramadolor or tramadura or tramagetic or tramagit or tramake or tramal or tramex or tramundin or trasedal or theradol or tiral or topalgic or tradol or tradolpuren or tradonal or tralgiol or tramadorsch or tramadin or tramadoc or ultram or zamudol or zumalgic or zydol or zytram).mp. |
| 47 | exp Animal Experiment/ |
| 48 | exp Experimental Animal/ |
| 49 | animal model/ |
| 50 | exp Rodent/ |
| 51 | (rat or rats or mouse or mice).ti. |
| 52 | longitudinal study/ |
| 53 | prospective study/ |
| 54 | retrospective study/ |
| 55 | cohort analysis/ |
| 56 | ((cohort or follow up or observational or epidemiologic*) adj (study or studies)).mp. |
| 57 | observational study/ |
| 60 | (Randomized controlled trial/ or Controlled clinical study/ or random*.ti,ab. or randomization/ or intermethod comparison/ or placebo.ti,ab. or (compare or compared or comparison).ti. or ((evaluated or evaluate or evaluating or assessed or assess) and (compare or compared or comparing or comparison)).ab. or (open adj label).ti,ab. or ((double or single or doubly or singly) adj (blind or blinded or blindly)).ti,ab. or double blind procedure/ or parallel group*1.ti,ab. or (crossover or cross over).ti,ab. or ((assign* or match or matched or allocation) adj5 (alternate or group*1 or intervention*1 or patient*1 or subject*1 or participant*1)).ti,ab. or (assigned or allocated).ti,ab. or (controlled adj7 (study or design or trial)).ti,ab. or (volunteer or volunteers).ti,ab. or human experiment/ or trial.ti.) not (((random* adj sampl* adj7 ("cross section*" or questionnaire*1 or survey* or database*1)).ti,ab. not (comparative study/ or controlled study/ or randomi?ed controlled.ti,ab. or randomly assigned.ti,ab.)) or (Cross-sectional study/ not (randomized controlled trial/ or controlled clinical study/ or controlled study/ or randomi?ed controlled.ti,ab. or control group*1.ti,ab.)) or (((case adj control*) and random*) not randomi?ed controlled).ti,ab. or (Systematic review not (trial or study)).ti. or (nonrandom* not random*).ti,ab. or "Random field*".ti,ab. or (random cluster adj3 sampl*).ti,ab. or ((review.ab. and review.pt.) not trial.ti.) or ("we searched".ab. and (review.ti. or review.pt.)) or "update review".ab. or (databases adj4 searched).ab. or ((rat or rats or mouse or mice or swine or porcine or murine or sheep or lambs or pigs or piglets or rabbit or rabbits or cat or cats or dog or dogs or cattle or bovine or monkey or monkeys or trout or marmoset*1).ti. and animal experiment/) or (Animal experiment/ not (human experiment/ or human/))) |
| 73 | exp smoking/ or smok*.mp. |

**Embase**

| 1 | Chronic Pain/ |
| --- | --- |
| 2 | (chronic adj4 pain*).mp. [mp=title, abstract, heading word, drug trade name, original title, device manufacturer, drug manufacturer, device trade name, keyword heading word, floating subheading word, candidate term word] |
| 3 | exp Osteoarthritis/ |
| 4 | osteo-arthritis.mp. |
| 5 | osteoarthr*.mp. [mp=title, abstract, heading word, drug trade name, original title, device manufacturer, drug manufacturer, device trade name, keyword heading word, floating subheading word, candidate term word] |
| 6 | degenerative arthrit*.mp. |
| 7 | exp rheumatoid arthritis/ |
| 8 | osteo?arthrit*.mp. |
| 9 | exp Neuralgia/ |
| 10 | Diabetic Neuropathies.mp. or Diabetic Neuropathies/ |
| 11 | (neuropath* adj5 (pain* or diabet*)).mp. |
| 12 | Neuralgia/ or neuralg*.mp. |
| 13 | fibromyalg*.mp. |
| 14 | ((noncancer* or non-cancer*or chronic* or recurrent or persist* or non-malign* or long term or longterm or wide spread or wide spread chronic) adj3 pain).mp. |
| 15 | (complex regional pain syndromes or causalgia).mp. |
| 16 | Hyperalgesia/ |
| 17 | Trigeminal Neuralgia/ or nerve neuralgia.mp. |
| 18 | (acute or emergency or cancer pain or pr preoperative or postoperative or palliative care or surgical).ti,ab. |
| 19 | deprescrib*.mp. |
| 20 | de-prescrib*.mp. |
| 21 | deprescription*.mp. |
| 22 | de-prescription*.mp. |
| 23 | discontin*.mp. |
| 24 | taper*.mp. |
| 25 | wean*.mp. |
| 26 | switch*.mp. |
| 27 | cess*.mp. |
| 28 | terminat*.mp. |
| 29 | substitut*.mp. |
| 30 | reflex sympathetic dystrophy.mp. |
| 31 | phantom limb.mp. or agnosia/ or phantom pain/ or amputation stump/ |
| 32 | exp backache/ |
| 33 | radiculopathy.mp. or exp radiculopathy/ |
| 34 | musculoskeletal pain/ |
| 35 | exp arthralgia/ |
| 36 | headache/ |
| 37 | headache*.mp. |
| 38 | exp migraine/ |
| 39 | migraine.mp. |
| 40 | fibromyalgia/ |
| 42 | exp narcotic analgesic agent/ |
| 43 | (opioid* or opiate*).mp. |
| 44 | (alfentanil or alphaprodine or beta-casomorphin$ or buprenorphine or carfentanil or codeine or deltorphin or dextromethorphan or dezocine or dihydrocodeine or dihydromorphine or enkephalin$ or ethylketocyclazocine or ethylmorphine or etorphine or fentanyl or heroin or hydrocodone or hydromorphone or ketobemidone or levorphanol or lofentanil or meperidine or meptazinol or methadone or methadyl acetate or morphine or nalbuphine or opium or oxycodone or oxymorphone or pentazocine or phenazocine or phenoperidine or pirinitramide or promedol or propoxyphene or remifentanil or sufentanil or tilidine or tapentadol).mp. |
| 45 | (adolonta or Anpec or Ardinex or Asimadoline or Alvimopam or amadol or biodalgic or biokanol or Codinovo or contramal or Demerol or Dicodid or Dihydrocodeinone or dihydromorphinone or dihydrohydroxycodeinone or dihydrone or dilaudid or dinarkon or dolsin or dolosal or dolin or dolantin or dolargan or dolcontral or duramorph or duromorph or duragesic or durogesic or eucodal or Fedotzine or Fentanest or Fentora or Fortral or Hycodan or Hycon or Hydrocodone or Hydrocodeinonebitartrate or hydromorphon or hydroxycodeinon or isocodeine or isonipecain or jutadol or laudacon or l dromoran or levodroman or levorphan or levo-dromoran or levodromoran or lexir or lidol or lydol or morfin or morfine or morphia or morphin or morphinium or morphinene or morphium or ms contin or n-methylmorphine or n methylmorphine or nobligan or numorphan or oramorph or oxycodeinon or oxiconum or oxycone or oxycontin or palladone or pancodine or pethidine or phentanyl or prontofort or robidone or skenan or sublimaze or sulfentanyl or sulfentanil or sufenta or takadol or talwin or theocodin or tramadol or tramadolhameln or tramadolor or tramadura or tramagetic or tramagit or tramake or tramal or tramex or tramundin or trasedal or theradol or tiral or topalgic or tradol or tradolpuren or tradonal or tralgiol or tramadorsch or tramadin or tramadoc or ultram or zamudol or zumalgic or zydol or zytram).mp. |
| 47 | exp Animal Experiment/ |
| 48 | exp Experimental Animal/ |
| 49 | animal model/ |
| 50 | exp Rodent/ |
| 51 | (rat or rats or mouse or mice).ti. |
| 52 | longitudinal study/ |
| 53 | prospective study/ |
| 54 | retrospective study/ |
| 55 | cohort analysis/ |
| 56 | ((cohort or follow up or observational or epidemiologic*) adj (study or studies)).mp. |
| 57 | observational study/ |
| 60 | (Randomized controlled trial/ or Controlled clinical study/ or random*.ti,ab. or randomization/ or intermethod comparison/ or placebo.ti,ab. or (compare or compared or comparison).ti. or ((evaluated or evaluate or evaluating or assessed or assess) and (compare or compared or comparing or comparison)).ab. or (open adj label).ti,ab. or ((double or single or doubly or singly) adj (blind or blinded or blindly)).ti,ab. or double blind procedure/ or parallel group*1.ti,ab. or (crossover or cross over).ti,ab. or ((assign* or match or matched or allocation) adj5 (alternate or group*1 or intervention*1 or patient*1 or subject*1 or participant*1)).ti,ab. or (assigned or allocated).ti,ab. or (controlled adj7 (study or design or trial)).ti,ab. or (volunteer or volunteers).ti,ab. or human experiment/ or trial.ti.) not (((random* adj sampl* adj7 ("cross section*" or questionnaire*1 or survey* or database*1)).ti,ab. not (comparative study/ or controlled study/ or randomi?ed controlled.ti,ab. or randomly assigned.ti,ab.)) or (Cross-sectional study/ not (randomized controlled trial/ or controlled clinical study/ or controlled study/ or randomi?ed controlled.ti,ab. or control group*1.ti,ab.)) or (((case adj control*) and random*) not randomi?ed controlled).ti,ab. or (Systematic review not (trial or study)).ti. or (nonrandom* not random*).ti,ab. or "Random field*".ti,ab. or (random cluster adj3 sampl*).ti,ab. or ((review.ab. and review.pt.) not trial.ti.) or ("we searched".ab. and (review.ti. or review.pt.)) or "update review".ab. or (databases adj4 searched).ab. or ((rat or rats or mouse or mice or swine or porcine or murine or sheep or lambs or pigs or piglets or rabbit or rabbits or cat or cats or dog or dogs or cattle or bovine or monkey or monkeys or trout or marmoset*1).ti. and animal experiment/) or (Animal experiment/ not (human experiment/ or human/))) |
| 73 | exp smoking/ or smok*.mp. |

Cochrane Library

| ID | Search |
| --- | --- |
| #1 | chronic near/3 pain |
| #2 | MeSH descriptor: [Chronic Pain] explode all trees |
| #3 | MeSH descriptor: [Osteoarthritis] explode all trees |
| #4 | osteoarthrit* |
| #5 | osteo-arthritis |
| #6 | degenerative arthrit* |
| #7 | MeSH descriptor: [Arthritis, Rheumatoid] explode all trees |
| #8 | MeSH descriptor: [Neuralgia] explode all trees |
| #9 | MeSH descriptor: [Diabetic Neuropathies] explode all trees |
| #10 | neuropath* near/5 (pain* or diabet*) |
| #11 | neuralg* |
| #12 | MeSH descriptor: [Migraine Disorders] explode all trees |
| #13 | migraine |
| #14 | MeSH descriptor: [Fibromyalgia] explode all trees |
| #15 | fibromyalg* |
| #16 | MeSH descriptor: [Complex Regional Pain Syndromes] explode all trees |
| #17 | complex regional pain syndromes or causalgia |
| #18 | MeSH descriptor: [Phantom Limb] explode all trees |
| #19 | ((noncancer* or non-cancer*or chronic* or recurrent or persist* or non-malign* or long term or long-term or longterm or widespread chronic) near/3 pain) |
| #20 | MeSH descriptor: [Hyperalgesia] explode all trees |
| #22 | MeSH descriptor: [Radiculopathy] explode all trees |
| #23 | MeSH descriptor: [Musculoskeletal Pain] explode all trees |
| #24 | MeSH descriptor: [Arthralgia] explode all trees |
| #25 | MeSH descriptor: [Headache Disorders] explode all trees |
| #26 | MeSH descriptor: [Headache] explode all trees |
| #27 | headache* |
| #28 | backache* or backpain* or dorsalgi* or arthralgi* or polyarthralgi* or arthrodyni* or myalgi* or fibromyalgi* or myodyni* or neuralgi* or ischialgi* or crps or rachialgi* |
| #29 | ((back or discogen* or bone or musculoskelet* or muscle* or skelet* or spinal or spine or vertebra* or joint* or arthritis or Intestin* or neuropath* or neck or cervical* or head or facial* or complex or radicular or cervicobrachi* or orofacial or somatic or shoulder* or knee* or hip or hips) near/3 pain) |
| #30 | radiculopathy |
| #31 | MeSH descriptor: [Back Pain] explode all trees |
| #33 | acute or emergency or preoperative or postoperative or surgical or cancer pain or palliative care |
| #36 | opioid* or opiate* |
| #37 | narcotic* |
| #38 | MeSH descriptor: [Analgesics, Opioid] explode all trees |
| #39 | MeSH descriptor: [Narcotics] explode all trees |
| #40 | alfentanil or alphaprodine or beta-casomorphin$ or buprenorphine or carfentanil or codeine or deltorphin or dextromethorphan or dezocine or dihydrocodeine or dihydromorphine or enkephalin$ or ethylketocyclazocine or ethylmorphine or etorphine or fentanyl or heroin or hydrocodone or hydromorphone or ketobemidone or levorphanol or lofentanil or meperidine or meptazinol or methadone or methadyl acetate or morphine or nalbuphine or opium or oxycodone or oxymorphone or pentazocine or phenazocine or phenoperidine or pirinitramide or promedol or propoxyphene or remifentanil or sufentanil or tilidine or tapentadol adolonta or Anpec or Ardinex or Asimadoline or Alvimopam or amadol or biodalgic or biokanol or Codinovo or contramal or Demerol or Dicodid or Dihydrocodeinone or dihydromorphinone or dihydrohydroxycodeinone or dihydrone or dilaudid or dinarkon or dolsin or dolosal or dolin or dolantin or dolargan or dolcontral or duramorph or duromorph or duragesic or durogesic or eucodal or Fedotzine or Fentanest or Fentora or Fortral or Hycodan or Hycon or Hydrocodone or Hydrocodeinonebitartrate or hydromorphon or hydroxycodeinon or isocodeine or isonipecain or jutadol or laudacon or l dromoran or levodroman or levorphan or levo-dromoran or levodromoran or lexir or lidol or lydol or morfin or morfine or morphia or morphin or morphinium or morphinene or morphium or ms contin or n-methylmorphine or n methylmorphine or nobligan or numorphan or oramorph or oxycodeinon or oxiconum or oxycone or oxycontin or palladone or pancodine or pethidine or phentanyl or prontofort or robidone or skenan or sublimaze or sulfentanyl or sulfentanil or sufenta or takadol or talwin or theocodin or tramadol or tramadolhameln or tramadolor or tramadura or tramagetic or tramagit or tramake or tramal or tramex or tramundin or trasedal or theradol or tiral or topalgic or tradol or tradolpuren or tradonal or tralgiol or tramadorsch or tramadin or tramadoc or ultram or zamudol or zumalgic or zydol or zytram |
| #44 | deprescrib* |
| #45 | de-prescrib* |
| #46 | deprescription* |
| #47 | de-prescription* |
| #48 | discontin* |
| #49 | taper* |
| #50 | wean* |
| #51 | switch* |
| #52 | cess* |
| #53 | terminat* |
| #54 | substitut* |
| #57 | animal* |
| #59 | child* |
| #60 | teenager* |
| #61 | Adolescent* |
| #62 | age under 18 |
| #63 | age below 18 |
| #64 | aged 18 and under |
| #65 | infant* |
| #66 | preschool* |
| #67 | pediatric* |
| #68 | toddler* |
| #69 | baby or babies |
| #70 | newborn |

**APA PsycInfo**

| 1 | chronic pain.mp. [mp=title, abstract, heading word, table of contents, key concepts, original title, tests & measures, mesh word] |
| --- | --- |
| 2 | chronic non cancer pain.mp. [mp=title, abstract, heading word, table of contents, key concepts, original title, tests & measures, mesh word] |
| 3 | chronic non-cancer pain.mp. [mp=title, abstract, heading word, table of contents, key concepts, original title, tests & measures, mesh word] |
| 4 | chronic noncancer pain.mp. [mp=title, abstract, heading word, table of contents, key concepts, original title, tests & measures, mesh word] |
| 5 | chronic non-malignant pain.mp. [mp=title, abstract, heading word, table of contents, key concepts, original title, tests & measures, mesh word] |
| 6 | chronic non malignant pain.mp. [mp=title, abstract, heading word, table of contents, key concepts, original title, tests & measures, mesh word] |
| 7 | chronic nonmalignant pain.mp. [mp=title, abstract, heading word, table of contents, key concepts, original title, tests & measures, mesh word] |
| 8 | long-term pain.mp. [mp=title, abstract, heading word, table of contents, key concepts, original title, tests & measures, mesh word] |
| 9 | long term pain.mp. [mp=title, abstract, heading word, table of contents, key concepts, original title, tests & measures, mesh word] |
| 10 | longterm pain.mp. [mp=title, abstract, heading word, table of contents, key concepts, original title, tests & measures, mesh word] |
| 11 | persistent pain*.mp. [mp=title, abstract, heading word, table of contents, key concepts, original title, tests & measures, mesh word] |
| 12 | chronic pain*.mp. [mp=title, abstract, heading word, table of contents, key concepts, original title, tests & measures, mesh word] |
| 13 | widespread pain*.mp. [mp=title, abstract, heading word, table of contents, key concepts, original title, tests & measures, mesh word] |
| 14 | widespread chronic pain*.mp. [mp=title, abstract, heading word, table of contents, key concepts, original title, tests & measures, mesh word] |
| 15 | lower back pain.mp. [mp=title, abstract, heading word, table of contents, key concepts, original title, tests & measures, mesh word] |
| 16 | low back pain.mp. [mp=title, abstract, heading word, table of contents, key concepts, original title, tests & measures, mesh word] |
| 17 | lower back ache.mp. [mp=title, abstract, heading word, table of contents, key concepts, original title, tests & measures, mesh word] |
| 18 | back ache.mp. [mp=title, abstract, heading word, table of contents, key concepts, original title, tests & measures, mesh word] |
| 19 | low back ache.mp. [mp=title, abstract, heading word, table of contents, key concepts, original title, tests & measures, mesh word] |
| 20 | postural low back pain.mp. [mp=title, abstract, heading word, table of contents, key concepts, original title, tests & measures, mesh word] |
| 21 | rheumatoid arthri*.mp. [mp=title, abstract, heading word, table of contents, key concepts, original title, tests & measures, mesh word] |
| 22 | osteoarthr*.mp. [mp=title, abstract, heading word, table of contents, key concepts, original title, tests & measures, mesh word] |
| 23 | degenerative arthritides.mp. [mp=title, abstract, heading word, table of contents, key concepts, original title, tests & measures, mesh word] |
| 24 | arthroses.mp. [mp=title, abstract, heading word, table of contents, key concepts, original title, tests & measures, mesh word] |
| 25 | osteoarthrosis deformans.mp. [mp=title, abstract, heading word, table of contents, key concepts, original title, tests & measures, mesh word] |
| 26 | musculoskeletal pain*.mp. [mp=title, abstract, heading word, table of contents, key concepts, original title, tests & measures, mesh word] |
| 27 | neuralgi*.mp. [mp=title, abstract, heading word, table of contents, key concepts, original title, tests & measures, mesh word] |
| 28 | neuropathic pain*.mp. [mp=title, abstract, heading word, table of contents, key concepts, original title, tests & measures, mesh word] |
| 29 | neurodynia.mp. [mp=title, abstract, heading word, table of contents, key concepts, original title, tests & measures, mesh word] |
| 30 | iliohypogastric nerve.mp. [mp=title, abstract, heading word, table of contents, key concepts, original title, tests & measures, mesh word] |
| 31 | nerve neuralgia.mp. [mp=title, abstract, heading word, table of contents, key concepts, original title, tests & measures, mesh word] |
| 32 | perineal neuralgia.mp. [mp=title, abstract, heading word, table of contents, key concepts, original title, tests & measures, mesh word] |
| 33 | supraorbital neuralgia.mp. [mp=title, abstract, heading word, table of contents, key concepts, original title, tests & measures, mesh word] |
| 34 | fibromyalgia.mp. [mp=title, abstract, heading word, table of contents, key concepts, original title, tests & measures, mesh word] |
| 35 | muscular rheumatism.mp. [mp=title, abstract, heading word, table of contents, key concepts, original title, tests & measures, mesh word] |
| 36 | myofascial pain syndrome.mp. [mp=title, abstract, heading word, table of contents, key concepts, original title, tests & measures, mesh word] |
| 37 | primary fibromyalgia.mp. [mp=title, abstract, heading word, table of contents, key concepts, original title, tests & measures, mesh word] |
| 38 | sciatic neuralgia.mp. [mp=title, abstract, heading word, table of contents, key concepts, original title, tests & measures, mesh word] |
| 39 | sciatic*.mp. [mp=title, abstract, heading word, table of contents, key concepts, original title, tests & measures, mesh word] |
| 40 | joint pain*.mp. [mp=title, abstract, heading word, table of contents, key concepts, original title, tests & measures, mesh word] |
| 41 | muscle pain*.mp. [mp=title, abstract, heading word, table of contents, key concepts, original title, tests & measures, mesh word] |
| 42 | muscle soreness.mp. [mp=title, abstract, heading word, table of contents, key concepts, original title, tests & measures, mesh word] |
| 43 | arthralgia*.mp. [mp=title, abstract, heading word, table of contents, key concepts, original title, tests & measures, mesh word] |
| 44 | muscle tenderness.mp. [mp=title, abstract, heading word, table of contents, key concepts, original title, tests & measures, mesh word] |
| 45 | neuralgi*.mp. [mp=title, abstract, heading word, table of contents, key concepts, original title, tests & measures, mesh word] |
| 46 | myalgi*.mp. [mp=title, abstract, heading word, table of contents, key concepts, original title, tests & measures, mesh word] |
| 47 | arthriti*.mp. [mp=title, abstract, heading word, table of contents, key concepts, original title, tests & measures, mesh word] |
| 48 | osteoarthr*.mp. [mp=title, abstract, heading word, table of contents, key concepts, original title, tests & measures, mesh word] |
| 49 | headache*.mp. [mp=title, abstract, heading word, table of contents, key concepts, original title, tests & measures, mesh word] |
| 50 | migrain*.mp. [mp=title, abstract, heading word, table of contents, key concepts, original title, tests & measures, mesh word] |
| 52 | analgesic*.mp. [mp=title, abstract, heading word, table of contents, key concepts, original title, tests & measures, mesh word] |
| 53 | opioid analgesic*.mp. [mp=title, abstract, heading word, table of contents, key concepts, original title, tests & measures, mesh word] |
| 54 | opioid*.mp. [mp=title, abstract, heading word, table of contents, key concepts, original title, tests & measures, mesh word] |
| 55 | opiate*.mp. [mp=title, abstract, heading word, table of contents, key concepts, original title, tests & measures, mesh word] |
| 56 | morphine*.mp. [mp=title, abstract, heading word, table of contents, key concepts, original title, tests & measures, mesh word] |
| 57 | codeine*.mp. [mp=title, abstract, heading word, table of contents, key concepts, original title, tests & measures, mesh word] |
| 58 | oxycodone*.mp. [mp=title, abstract, heading word, table of contents, key concepts, original title, tests & measures, mesh word] |
| 59 | tramadol*.mp. [mp=title, abstract, heading word, table of contents, key concepts, original title, tests & measures, mesh word] |
| 60 | fentanyl*.mp. [mp=title, abstract, heading word, table of contents, key concepts, original title, tests & measures, mesh word] |
| 61 | dihydrocodeine*.mp. [mp=title, abstract, heading word, table of contents, key concepts, original title, tests & measures, mesh word] |
| 62 | hydromorphone*.mp. [mp=title, abstract, heading word, table of contents, key concepts, original title, tests & measures, mesh word] |
| 63 | meperidine*.mp. [mp=title, abstract, heading word, table of contents, key concepts, original title, tests & measures, mesh word] |
| 64 | pethidine*.mp. [mp=title, abstract, heading word, table of contents, key concepts, original title, tests & measures, mesh word] |
| 65 | dextropropoxyphene*.mp. [mp=title, abstract, heading word, table of contents, key concepts, original title, tests & measures, mesh word] |
| 66 | pentazocine*.mp. [mp=title, abstract, heading word, table of contents, key concepts, original title, tests & measures, mesh word] |
| 67 | hydrocodone*.mp. [mp=title, abstract, heading word, table of contents, key concepts, original title, tests & measures, mesh word] |
| 68 | tapentadol*.mp. [mp=title, abstract, heading word, table of contents, key concepts, original title, tests & measures, mesh word] |
| 69 | diamorphine*.mp. [mp=title, abstract, heading word, table of contents, key concepts, original title, tests & measures, mesh word] |
| 70 | opium*.mp. [mp=title, abstract, heading word, table of contents, key concepts, original title, tests & measures, mesh word] |
| 71 | butorphanol*.mp. [mp=title, abstract, heading word, table of contents, key concepts, original title, tests & measures, mesh word] |
| 72 | diacetylmorphine*.mp. [mp=title, abstract, heading word, table of contents, key concepts, original title, tests & measures, mesh word] |
| 73 | heroin*.mp. [mp=title, abstract, heading word, table of contents, key concepts, original title, tests & measures, mesh word] |
| 74 | dextromethorphan*.mp. [mp=title, abstract, heading word, table of contents, key concepts, original title, tests & measures, mesh word] |
| 75 | naloxone*.mp. [mp=title, abstract, heading word, table of contents, key concepts, original title, tests & measures, mesh word] |
| 77 | deprescrib*.mp. [mp=title, abstract, heading word, table of contents, key concepts, original title, tests & measures, mesh word] |
| 78 | de-prescrib*.mp. [mp=title, abstract, heading word, table of contents, key concepts, original title, tests & measures, mesh word] |
| 79 | deprescription*.mp. [mp=title, abstract, heading word, table of contents, key concepts, original title, tests & measures, mesh word] |
| 80 | de-prescription*.mp. [mp=title, abstract, heading word, table of contents, key concepts, original title, tests & measures, mesh word] |
| 81 | discontin*.mp. [mp=title, abstract, heading word, table of contents, key concepts, original title, tests & measures, mesh word] |
| 82 | taper*.mp. [mp=title, abstract, heading word, table of contents, key concepts, original title, tests & measures, mesh word] |
| 83 | wean*.mp. [mp=title, abstract, heading word, table of contents, key concepts, original title, tests & measures, mesh word] |
| 84 | switch*.mp. [mp=title, abstract, heading word, table of contents, key concepts, original title, tests & measures, mesh word] |
| 85 | reduc*.mp. [mp=title, abstract, heading word, table of contents, key concepts, original title, tests & measures, mesh word] |
| 86 | cess*.mp. [mp=title, abstract, heading word, table of contents, key concepts, original title, tests & measures, mesh word] |
| 87 | terminat*.mp. [mp=title, abstract, heading word, table of contents, key concepts, original title, tests & measures, mesh word] |
| 88 | substitut*.mp. [mp=title, abstract, heading word, table of contents, key concepts, original title, tests & measures, mesh word] |
| 90 | adult patient*.mp. [mp=title, abstract, heading word, table of contents, key concepts, original title, tests & measures, mesh word] |
| 91 | human patient*.mp. [mp=title, abstract, heading word, table of contents, key concepts, original title, tests & measures, mesh word] |
| 92 | human people.mp. [mp=title, abstract, heading word, table of contents, key concepts, original title, tests & measures, mesh word] |
| 93 | human population*.mp. [mp=title, abstract, heading word, table of contents, key concepts, original title, tests & measures, mesh word] |
| 94 | adult people.mp. [mp=title, abstract, heading word, table of contents, key concepts, original title, tests & measures, mesh word] |
| 95 | adult population*.mp. [mp=title, abstract, heading word, table of contents, key concepts, original title, tests & measures, mesh word] |
| 96 | middle-aged patient*.mp. [mp=title, abstract, heading word, table of contents, key concepts, original title, tests & measures, mesh word] |
| 97 | middle aged patient*.mp. [mp=title, abstract, heading word, table of contents, key concepts, original title, tests & measures, mesh word] |
| 98 | middle-aged people.mp. [mp=title, abstract, heading word, table of contents, key concepts, original title, tests & measures, mesh word] |
| 99 | middle-aged population*.mp. [mp=title, abstract, heading word, table of contents, key concepts, original title, tests & measures, mesh word] |
| 100 | middle aged population*.mp. [mp=title, abstract, heading word, table of contents, key concepts, original title, tests & measures, mesh word] |
| 101 | elderly patient*.mp. [mp=title, abstract, heading word, table of contents, key concepts, original title, tests & measures, mesh word] |
| 102 | elderly population*.mp. [mp=title, abstract, heading word, table of contents, key concepts, original title, tests & measures, mesh word] |
| 103 | elderly people.mp. [mp=title, abstract, heading word, table of contents, key concepts, original title, tests & measures, mesh word] |
| 104 | (aged 18 or over).mp. [mp=title, abstract, heading word, table of contents, key concepts, original title, tests & measures, mesh word] |
| 105 | (aged 18 and over).mp. [mp=title, abstract, heading word, table of contents, key concepts, original title, tests & measures, mesh word] |
| 106 | (aged 65 and over).mp. [mp=title, abstract, heading word, table of contents, key concepts, original title, tests & measures, mesh word] |
| 107 | (aged 65 or over).mp. [mp=title, abstract, heading word, table of contents, key concepts, original title, tests & measures, mesh word] |
| 108 | (aged 65 and older).mp. [mp=title, abstract, heading word, table of contents, key concepts, original title, tests & measures, mesh word] |
| 109 | (aged 65 or older).mp. [mp=title, abstract, heading word, table of contents, key concepts, original title, tests & measures, mesh word] |
| 110 | (aged 18 and older).mp. [mp=title, abstract, heading word, table of contents, key concepts, original title, tests & measures, mesh word] |
| 111 | (aged 18 or older).mp. [mp=title, abstract, heading word, table of contents, key concepts, original title, tests & measures, mesh word] |
| 112 | 18-65.mp. [mp=title, abstract, heading word, table of contents, key concepts, original title, tests & measures, mesh word] |
| 113 | between 18.mp. [mp=title, abstract, heading word, table of contents, key concepts, original title, tests & measures, mesh word] |
| 114 | from 18 to 65.mp. [mp=title, abstract, heading word, table of contents, key concepts, original title, tests & measures, mesh word] |
| 117 | palliative pain*.mp. [mp=title, abstract, heading word, table of contents, key concepts, original title, tests & measures, mesh word] |
| 118 | cancer pain*.mp. [mp=title, abstract, heading word, table of contents, key concepts, original title, tests & measures, mesh word] |
| 119 | acute pain*.mp. [mp=title, abstract, heading word, table of contents, key concepts, original title, tests & measures, mesh word] |
| 120 | surgical pain*.mp. [mp=title, abstract, heading word, table of contents, key concepts, original title, tests & measures, mesh word] |
| 124 | randomised controlled trial*.mp. [mp=title, abstract, heading word, table of contents, key concepts, original title, tests & measures, mesh word] |
| 125 | randomized controlled trial*.mp. [mp=title, abstract, heading word, table of contents, key concepts, original title, tests & measures, mesh word] |
| 126 | controlled clinical trial*.mp. [mp=title, abstract, heading word, table of contents, key concepts, original title, tests & measures, mesh word] |
| 127 | randomized.mp. [mp=title, abstract, heading word, table of contents, key concepts, original title, tests & measures, mesh word] |
| 128 | randomised.mp. [mp=title, abstract, heading word, table of contents, key concepts, original title, tests & measures, mesh word] |
| 129 | clinical trial*.mp. [mp=title, abstract, heading word, table of contents, key concepts, original title, tests & measures, mesh word] |
| 130 | placebo*.mp. [mp=title, abstract, heading word, table of contents, key concepts, original title, tests & measures, mesh word] |
| 131 | randomisation*.mp. [mp=title, abstract, heading word, table of contents, key concepts, original title, tests & measures, mesh word] |
| 132 | randomization*.mp. [mp=title, abstract, heading word, table of contents, key concepts, original title, tests & measures, mesh word] |
| 133 | phase 1.mp. [mp=title, abstract, heading word, table of contents, key concepts, original title, tests & measures, mesh word] |
| 134 | phase I.mp. [mp=title, abstract, heading word, table of contents, key concepts, original title, tests & measures, mesh word] |
| 135 | phase one.mp. [mp=title, abstract, heading word, table of contents, key concepts, original title, tests & measures, mesh word] |
| 136 | phase 2.mp. [mp=title, abstract, heading word, table of contents, key concepts, original title, tests & measures, mesh word] |
| 137 | phase II.mp. [mp=title, abstract, heading word, table of contents, key concepts, original title, tests & measures, mesh word] |
| 138 | phase two.mp. [mp=title, abstract, heading word, table of contents, key concepts, original title, tests & measures, mesh word] |
| 139 | phase 3.mp. [mp=title, abstract, heading word, table of contents, key concepts, original title, tests & measures, mesh word] |
| 140 | phase III.mp. [mp=title, abstract, heading word, table of contents, key concepts, original title, tests & measures, mesh word] |
| 141 | phase three.mp. [mp=title, abstract, heading word, table of contents, key concepts, original title, tests & measures, mesh word] |
| 142 | phase 4.mp. [mp=title, abstract, heading word, table of contents, key concepts, original title, tests & measures, mesh word] |
| 143 | phase IV.mp. [mp=title, abstract, heading word, table of contents, key concepts, original title, tests & measures, mesh word] |
| 144 | phase four.mp. [mp=title, abstract, heading word, table of contents, key concepts, original title, tests & measures, mesh word] |
| 145 | single-blind.mp. [mp=title, abstract, heading word, table of contents, key concepts, original title, tests & measures, mesh word] |
| 146 | double-blind.mp. [mp=title, abstract, heading word, table of contents, key concepts, original title, tests & measures, mesh word] |
| 147 | triple-blind.mp. [mp=title, abstract, heading word, table of contents, key concepts, original title, tests & measures, mesh word] |
| 149 | cohort stud*.mp. [mp=title, abstract, heading word, table of contents, key concepts, original title, tests & measures, mesh word] |
| 150 | prospective cohort stud*.mp. [mp=title, abstract, heading word, table of contents, key concepts, original title, tests & measures, mesh word] |
| 151 | retrospective cohort stud*.mp. [mp=title, abstract, heading word, table of contents, key concepts, original title, tests & measures, mesh word] |
| 152 | cohort analys*.mp. [mp=title, abstract, heading word, table of contents, key concepts, original title, tests & measures, mesh word] |
| 153 | historical cohort stud*.mp. [mp=title, abstract, heading word, table of contents, key concepts, original title, tests & measures, mesh word] |

CINAIL

| 1 | (MH "Chronic Pain") |
| --- | --- |
| 2 | chronic N4 pain |
| 3 | (MH "Osteoarthritis+") |
| 4 | "osteoarthrit*" |
| 5 | "osteo-arthritis" |
| 6 | "degenerative arthrit*" |
| 7 | (MH "Arthritis, Rheumatoid+") |
| 8 | (MH "Neuralgia+") |
| 9 | (MH "Diabetic Neuropathies") |
| 10 | (neuropath* N5 (pain* or diabet*)) |
| 11 | "neuralg*" |
| 12 | (MH "Migraine") |
| 13 | "migraine" |
| 14 | (MH "Fibromyalgia") |
| 15 | "fibromyalg*" |
| 16 | (MH "Complex Regional Pain Syndromes+") |
| 17 | "causalgia" |
| 18 | (MH "Phantom Limb") OR (MH "Phantom Pain") |
| 19 | (MH "Hyperalgesia") |
| 20 | ((noncancer* or non-cancer*or chronic* or recurrent or persist* or non-malign*) N3 pain) |
| 22 | (MH "Back Pain+") |
| 23 | (MH "Radiculopathy") OR "radiculopathy" |
| 24 | "musculoskeletal pain" |
| 25 | (MH "Arthralgia+") |
| 26 | (MH "Headache") |
| 27 | "headache*" |
| 28 | backache* or backpain* or dorsalgi* or arthralgi* or polyarthralgi* or arthrodyni* or myalgi* or fibromyalgi* or myodyni* or neuralgi* or ischialgi* or crps or rachialgi* |
| 29 | ((back or discogen* or bone or musculoskelet* or muscle* or skelet* or spinal or spine or vertebra* or joint* or arthritis or Intestin* or neuropath* or neck or cervical* or head or facial* or complex or radicular or cervicobrachi* or orofacial or somatic or shoulder* or knee* or hip or hips) N3 pain) |
| 31 | acute or emergency or preoperative or postoperative or surgical or palliative care or cancer pain |
| 34 | (MH "Analgesics, Opioid+") |
| 35 | "opioid*" |
| 36 | "opiate*" |
| 37 | (MH "Narcotics+") |
| 38 | "narcotic*" |
| 39 | alfentanil or alphaprodine or beta-casomorphin$ or buprenorphine or carfentanil or codeine or deltorphin or dextromethorphan or dezocine or dihydrocodeine or dihydromorphine or enkephalin$ or ethylketocyclazocine or ethylmorphine or etorphine or fentanyl or heroin or hydrocodone or hydromorphone or ketobemidone or levorphanol or lofentanil or meperidine or meptazinol or methadone or methadyl acetate or morphine or nalbuphine or opium or oxycodone or oxymorphone or pentazocine or phenazocine or phenoperidine or pirinitramide or promedol or propoxyphene or remifentanil or sufentanil or tilidine or tapentadol |
| 40 | adolonta or Anpec or Ardinex or Asimadoline or Alvimopam or amadol or biodalgic or biokanol or Codinovo or contramal or Demerol or Dicodid or Dihydrocodeinone or dihydromorphinone or dihydrohydroxycodeinone or dihydrone or dilaudid or dinarkon or dolsin or dolosal or dolin or dolantin or dolargan or dolcontral or duramorph or duromorph or duragesic or durogesic or eucodal or Fedotzine or Fentanest or Fentora or Fortral or Hycodan or Hycon or Hydrocodone or Hydrocodeinonebitartrate or hydromorphon or hydroxycodeinon or isocodeine or isonipecain or jutadol or laudacon or l dromoran or levodroman or levorphan or levo-dromoran or levodromoran or lexir or lidol or lydol or morfin or morfine or morphia or morphin or morphinium or morphinene or morphium or ms contin or n-methylmorphine or n-methylmorphine or nobligan or numorphan or oramorph or oxycodeinon or oxiconum or oxycone or oxycontin or palladone or pancodine or pethidine or phentanyl or prontofort or robidone or skenan or sublimaze or sulfentanyl or sulfentanil or sufenta or takadol or talwin or theocodin or tramadol or tramadolhameln or tramadol or tramadura or tramagetic or tramagit or tramake or tramal or tramex or tramundin or trasedal or theradol or tiral or topalgic or tradol or tradolpuren or tradonal or tralgiol or tramadorsch or tramadin or tramadoc or ultram or zamudol or zumalgic or zydol or zytram |
| 43 | deprescrib* |
| 44 | de-prescrib* |
| 45 | deprescription* |
| 46 | de-prescription* |
| 47 | discontin* |
| 48 | taper* |
| 49 | wean* |
| 50 | switch* |
| 51 | cess* |
| 52 | terminat* |
| 53 | substitut* |
| 56 | (MH "Prospective Studies+") |
| 57 | (MH "Correlational Studies") |
| 58 | (cohort N2 (study or studies)) |
| 59 | (observational N2 (study or studies)) |
| 60 | longitudinal studies or longitudinal research or longitudinal method |
| 61 | follow-up study |
| 62 | retrospective cohort study |
| 63 | prospective cohort study |
| 64 | cohort analy* |
| 65 | before and after comparison |
| 66 | before-after comparison |
| 67 | epidemiological study |
| 69 | (MH "Animals+") |
| 70 | S68 NOT S69 |
| 71 | (MH "Clinical Trials+") |
| 72 | clinical W3 trial |
| 73 | "double-blind" |
| 74 | "single-blind" |
| 75 | "triple-blind" |
| 77 | (MH "Placebo Effect") |
| 78 | (MH "Placebos") |
| 79 | "placebo*" |
| 80 | "random*" |
| 82 | (MH "Random Sample+") |
| 83 | (MH "Study Design") OR (MH "Crossover Design") OR (MH "Experimental Studies+") |
| 84 | "latin square" |
| 85 | (MH "Comparative Studies") |
| 86 | (MH "Evaluation Research+") |
| 87 | (MH "Prospective Studies+") |
| 90 | (MH "Animals+") |
